# Heterogeneity in Lipoprotein(a) Profile Changes Across the Menopausal Transition

**DOI:** 10.64898/2026.03.23.26349133

**Authors:** Catherine A. Palmer, Christy L. Avery, Christie M. Ballantyne, Misa Graff, Ron C. Hoogeveen, Anne Marie Jukic, Katherine M. Conners

## Abstract

**Introduction:** Menopause may coincide with rising Lp(a) levels, a causal risk factor for atherosclerotic cardiovascular disease (ASCVD). Characterizing changes in Lp(a) across menopause may inform risk stratification and testing recommendations.

**Methods:** We examined changes in serum Lp(a) levels by menopausal status among women with Lp(a) measured at visits 1 and 2 in the UK Biobank. Lp(a) analyses were examined by menopausal status: those who underwent menopause (N=415), those who remained premenopausal (N=532), and those who remained postmenopausal (N=3,615) between visits. We examined the change in Lp(a) between visits stratified by visit 1 Lp(a) levels. The primary outcome was incident Lp(a) ≥125 nmol/L at visit 2, estimated using Poisson regression with adjustment for baseline age.

**Results:** Data were available for 4,562 women (mean age at visit 1 = 57±7 years; median Lp(a) at visit 1 = 22 (IQR: 47) nmol/L; median time between visits = 4 (IQR: 1) years). At visit 1, median Lp(a) was slightly higher in postmenopausal women (23 nmol/L) than premenopausal women (19 nmol/L). Overall, median changes in Lp(a) between visits 1 and 2 were modest. Among women with intermediate visit 1 Lp(a) levels (75-125 nmol/L), those who transitioned through menopause experienced a median increase of 34.9 (−6.7, 53.0) nmol/L between visits, an approximately fourfold greater increase than for women who remained pre- (7.9 nmol/L) or postmenopausal (8.0 nmol/L). Further, 56% of women with intermediate visit 1 Lp(a) levels who transitioned through menopause between visits had incident Lp(a) ≥125 nmol/L at visit 2, compared with 29% and 28% of women who remained pre- or postmenopausal, representing an age-adjusted risk ratio of 2.26 (95% CI: 1.31, 3.90).

**Conclusion:** Relying on a single lifetime Lp(a) measurement may miss clinically relevant increases during menopause. Repeat testing in women as they age may improve identification of those at high risk for ASCVD.

**Clinical Perspective:** *What Is New?:*

- In this large longitudinal cohort, women who transitioned through menopause between visits experienced greater increases in Lp(a) than women who remained pre- or postmenopausal, particularly among those with intermediate baseline levels (75-125 nmol/L).
- More than half of women with intermediate Lp(a) who underwent menopause exceeded the ASCVD risk-enhancing threshold of 125 nmol/L.
- These findings provide quantitative estimates of menopause-associated reclassification across clinically relevant Lp(a) thresholds.

*What Are the Clinical Implications?:*

- A single premenopausal Lp(a) measurement may miss clinically meaningful increases that occur during the menopausal transition.
- Repeat Lp(a) testing after menopause may improve identification of women who newly meet risk-enhancing thresholds and may benefit from more intensive ASCVD risk assessment and prevention.
- Menopause should be considered a clinically important window for reassessment of Lp(a), particularly in women with intermediate premenopausal levels.

## Introduction

Cardiovascular disease (CVD) is the leading cause of death in women, yet is commonly underdiagnosed, in part because women are less likely than men to receive routine risk factor assessments.^1,2^ Prior to menopause, women experience a lower risk of CVD than men, however, their risk increases substantially during the menopausal transition, a period associated with adverse changes in lipid metabolism, blood pressure, and body fat distribution.^3,4^ A complete understanding of the mechanisms through which menopause may influence CVD risk remains elusive despite years of research, specifically whether menopause itself is a risk factor or if the observed increase in CVD risk is due to aging.

Lipoprotein (a) [Lp(a)] is a cholesterol-carrying lipoprotein that is an independent and causal atherosclerotic cardiovascular disease (ASCVD) risk factor with pro-atherogenic, pro-thrombotic, and pro-inflammatory properties.^5^ Although Lp(a) levels have been shown to remain relatively stable throughout the life course in men^6^, a small but growing body of evidence suggests that Lp(a) levels may increase in women around the time of menopause.^7–17^ Importantly, the degree to which Lp(a) changes across menopause remains unclear, as is whether such changes are large enough to result in clinically significant transitions across ASCVD risk-enhancing thresholds during the menopausal transition.

This uncertainty has direct clinical relevance. Current guidelines from the American College of Cardiology/American Heart Association (ACC/AHA) and the European Atherosclerosis Society (EAS) identify elevated Lp(a) as a risk-enhancing factor for ASCVD, and thresholds of 125 nmol/L (ACC/AHA) and 105 nmol/L (EAS) are increasingly used to guide preventive therapy.^18,19^ Although Lp(a) is often described as sufficiently stable to measure once in a lifetime, emerging guidance suggests that a second measurement after menopause may be clinically useful.^19,20^ Clarifying longitudinal patterns of Lp(a) during the menopausal transition is therefore essential for informing screening recommendations and ASCVD risk assessment in women.

To date, most studies examining menopause and Lp(a) have been cross-sectional, making it difficult to determine whether observed differences are attributable to the menopausal transition, to underlying age-related processes, or to population differences.^7–16^ The limited number of longitudinal studies to date have yielded inconsistent results, reflecting differences in study design and Lp(a) measurement methods, which complicate comparisons across studies.^6,17^

The objective of the present study is to assess longitudinal changes in Lp(a) levels and transitions across clinically meaningful ASCVD risk-thresholds in women with pre- and post-menopausal measurements in the UK Biobank.

## Methods

### Study Population

The UK Biobank is a large prospective cohort study that enrolled over 500,000 individuals aged 40 to 69 years old between 2006 and 2010.^21^ Eligible adults registered with the UK’s National Health Service were invited to attend a baseline visit (visit 1) at one of 22 assessment centers.

During the baseline assessment, detailed information on lifestyle and medical history was collected through a touchscreen questionnaire and face-to-face interviews, followed by physical measurements and biological samples. Participants living within 25 miles of the UK Biobank coordinating center in Stockport, UK (n=20,000) were invited to undergo a follow-up visit (visit 2) between 2012 and 2013.^21^ Women were eligible for the present analysis if they had self-reported menopausal status at both visits and measured Lp(a) within the reportable range (3.8-189 nmol/L) at both visits. Given the very small number of participants who identified as non-European ancestry (<2%), and the consequent inability to generate stable population-specific estimates, these women were excluded from the analytic cohort. Women who reported themselves as postmenopausal at visit 1 and premenopausal at visit 2 were also excluded (n=11).

### Measurements

Serum Lp(a) levels were measured using isoform insensitive immunoturbidimetry on a Beckman Coulter AU5800 (nmol/L) (Randox Bioscience, UK).^22^ Menopausal status was self-reported at visit 1 and visit 2. Age was derived by the UK Biobank from birthdays of participants and the date they attended the assessment center, truncated to the whole year. To characterize the cohort at visit 1, we described the following covariates. BMI was calculated by the UK Biobank from height and weight measured at the assessment center visit. Age at menopause, use of hormone-replacement therapy (HRT), and smoking status were self-reported. Diabetes was determined via self-report, diabetes medication use, or HbA1c >6.5%, and hypertension was determined via self-report, antihypertensive medication use, or systolic blood pressure >140/90 mmHg. eGFR was estimated via the 2021 CKD-EPI equation. Low density lipoprotein cholesterol (LDL-c) levels were measured using the Nightingale Health NMR biomarker platform (Nightingale Health Plc.) with 500 MHz NMR spectrometers (Bruker AVANCE IIIHD).^23^

### Statistical Analyses

Visit 1 characteristics were summarized overall and by menopausal trajectory: women who remained premenopausal, women who transitioned through menopause, and women who remained postmenopausal. Continuous variables were summarized using means (SD) or medians (IQR), as appropriate, and categorical variables were summarized using frequencies and percentages.

Changes in Lp(a) between visit 1 and visit 2 were examined overall, within each menopausal trajectory group, and across clinically defined visit 1 Lp(a) categories. Absolute changes in continuous Lp(a) between visits were compared across menopausal trajectory groups using general linear models. Lp(a) was also modeled categorically according to the ACC/AHA risk-enhancing threshold of ≥125 nmol/L.^18^ We further characterized <75 nmol/L as low and 75-125 nmol/L as intermediate Lp(a) based on the classification of risk levels identified by the National Lipid Association (NLA).^20^ McNemar’s test was used to assess within-person change across the 125 nmol/L threshold. Among women with Lp(a) <125 nmol/L at visit 1, we estimated risk ratios for transitioning to elevated Lp(a) (≥125 nmol/L) at visit 2 using Poisson regression with a log link, modeling menopausal transition status as the primary exposure and adjusting for baseline age. No further adjustments were made because Lp(a) is generally invariant to lifestyle and clinical factors. Because women with Lp(a) levels in the intermediate range (75-125 nmol/L) are most plausibly at risk of crossing established risk-enhancing thresholds, analyses of incident elevated Lp(a) were focused on this subgroup to improve clinical interpretability. Analyses were repeated using 105 nmol/L as the risk-enhancing threshold. All analyses were performed in SAS version 9.4 (SAS Institute, Cary, NC).

## Results

### Participant Characteristics

Among 4,562 women who attended the visit 1 and visit 2 UK Biobank exams (mean age at visit 1 = 57±7 years, median time between visits = 4 years), 532 (12%) remained premenopausal at visit 2, 3,615 (79%) were postmenopausal at both visits, and 415 (9%) transitioned through menopause between visits. Women who were postmenopausal at visit 1 had higher LDL-C levels and higher prevalences of hypertension and diabetes at visit 1 compared to women who were premenopausal at both visits and women who underwent menopause between visits (Table 1). Median Lp(a) at visit 1 was 18.6 nmol/L for premenopausal women and slightly higher for postmenopausal women (23.0 nmol/L). At visit 2, the median time since menopause was 2 years for women who had menopause between visits, compared to 13 years for women who were postmenopausal at both visits.

**Table 1.** Participant Characteristics at Visit 1 by Menopausal Transition Status. Values are mean (SD) or N (%) unless stated otherwise.

| **Characteristic** | **Overall**<br>**(N = 4,562)** | **Women who**<br>**remained pre-**<br>**menopausal**<br>**(N = 532)** | **Women who**<br>**remained post-**<br>**menopausal**<br>**(N = 3,615)** | **Women who**<br>**underwent**<br>**menopause**<br>**(N = 415)** |
| --- | --- | --- | --- | --- |
| Age, years | 57.19 (7.24) | 44.22 (2.97) | 59.97 (4.93) | 49.65 (3.68) |
| Lp(a), (nmol/L) median (25<sup>th</sup> percentile, 75<sup>th</sup> percentile) | 22.10<br>(10.20, 55.94) | 19.40<br>(9.60, 58.45) | 23.06<br>(10.40, 56.43) | 16.59<br>(8.49, 48.5) |
| BMI<sup>a</sup> | 26.19 (4.71) | 25.67 (4.90) | 26.31 (4.67) | 25.86 (4.79) |
| Prevalent hypertension | 1930 (44.41) | 90 (17.54) | 1705 (49.51) | 135 (34.70) |
| Current smoker | 238 (5.22) | 33 (6.20) | 170 (4.70) | 35 (8.43) |
| Prevalent diabetes<sup>c</sup> | 121 (2.66) | 5 (0.95) | 106 (2.94) | 10 (2.42) |
| Current hormone replacement therapy use | 224 (4.91) | 1 (0.19) | 208 (5.75) | 15 (3.61) |
| eGFR (ml/min/1.73 m<sup>2</sup>) | 96.50 (13.35) | 107.17 (10.20) | 94.00 (12.83) | 104.63 (11.26) |
| LDL-C (mg/dL) | 139.71 (32.26) | 123.72 (27.68) | 143.21 (32.33) | 129.73 (28.71) |

<sup>a</sup>N missing for BMI = 80
<sup>b</sup>N missing for prevalent hypertension = 216
<sup>c</sup>N missing for prevalent diabetes = 14

### Lp(a) Transitions Between Visits

Across the entire study population, Lp(a) did not meaningfully change between visits (median change +0.9 nmol/L [-2.6, 6.8]) (Table 2). When stratified by menopausal transition status, women who transitioned through menopause experienced a slightly larger median increase between visits (+3.0 [–0.7, 11.2] nmol/L), compared to women who remained premenopausal (+0.7 [–2.6, 7.2] nmol/L) and women who were postmenopausal (+0.7 [–2.9, 6.3] nmol/L).

**Table 2.** Lp(a) at visit 1 and visit 2 by menopausal transition status (nmol/L). Values are expressed as median (25^th^ percentile, 75^th^ percentile)

|  | **Median**<br>**Lp(a) at visit 1** | **Median**<br>**Lp(a) at visit 2** | **Median change in Lp(a)**<br>**between visits** |
| --- | --- | --- | --- |
| **Overall (N=4,562)** | 22.1<br>(10.2, 55.9) | 23.6<br>(10.8, 58.9) | 0.9<br>(-2.6, 6.8) |
| **Women who underwent**<br>**menopause (N=415)** | 16.6<br>(8.5, 48.5) | 22.7<br>(10.9, 55.3) | 3.0<br>(-0.7, 11.2) |
| **Women who remained pre-**<br>**menopausal (N=532)** | 19.4<br>(9.6, 58.5) | 21.2<br>(9.9, 62.2) | 0.7<br>(-2.6, 7.2) |
| **Women who remained**<br>**post-menopausal (N=3,615)** | 23.1<br>(10.4, 56.4) | 23.9<br>(11.1, 59.1) | 0.7<br>(-2.9, 6.3) |

However, when stratified by visit 1 Lp(a), pronounced changes across menopause were observed, particularly among women with intermediate Lp(a) levels (75-125 nmol/L). In this subgroup, women who transitioned through menopause (n=22) experienced a median increase of 34.9 nmol/L between visits, approximately fourfold larger than women who remained premenopausal (n=20, +7.9 nmol/L) or postmenopausal (n=89, +8.0 nmol/L). Women who underwent menopause between visits also exhibited the largest median change in Lp(a) among individuals with low visit 1 levels (<75 nmol/L) and among those with elevated visit 1 levels (≥125 nmol/L), although these changes were modest (Table 3).

**Table 3.** Changes in Lp(a) between visit 1 and visit 2 by menopausal transition status and visit 1 Lp(a) level (nmol/L). Values are expressed as median (25^th^ percentile, 75^th^ percentile).

|  | <75 nmol/L at visit 1 | 75-125 nmol/L at visit 1 | ≥125 nmol/L at visit 1 |
| --- | --- | --- | --- |
| **Overall**<br>(N = 4,562) | 0.8<br>(-2.0, 5.1)<br>(n=3,708) | 9.3<br>(-6.5, 30.4)<br>(n=426) | -1.3<br>(-16.6, 16.2)<br>(n=428) |
| **Women who underwent menopause**<br>(N = 415) | 2.5<br>(-0.5, 8.4)<br>(n=348) | 34.9<br>(-6.7, 53.0)<br>(n=39) | 6.4<br>(-13.9, 23.4)<br>(n=28) |
| **Women who remained premenopausal**<br>(N = 532) | 0.6<br>(-1.9, 4.8)<br>(n=427) | 7.9<br>(-6.6, 24.2)<br>(n=69) | -2.2<br>(-27.5, 17.1)<br>(n=36) |
| **Women who remained postmenopausal**<br>(N = 3,615) | 0.7<br>(-2.3, 4.7)<br>(n=2,933) | 8.0<br>(-6.5, 27.7)<br>(n=318) | -1.5<br>(-16.5, 15.7)<br>(n=364) |

### Changes Across Risk Thresholds

A higher percentage of women who underwent menopause exceeded the risk-enhancing threshold of 125 nmol/L at visit 2 than those in the pre- and post-menopausal groups. This difference was particularly pronounced among women with intermediate Lp(a) levels at visit 1 (75-125 nmol/L). In this subgroup, more than half (56.4%) of women who underwent menopause exceeded 125 nmol/L at visit 2, nearly double the proportion observed among women who remained premenopausal (29.0%) or postmenopausal (28.0%) (Table 4). Among women with intermediate Lp(a) levels at visit 1 (75-125 nmol/L), those who transitioned through menopause between visits had more than twice the risk of developing elevated Lp(a) by visit 2 compared with women who remained premenopausal after adjustment for age at visit 1 (risk ratio [RR]: 2.08; 95% CI: 1.28-3.36). A similarly elevated risk was observed when women who transitioned through menopause were compared with those who remained premenopausal or postmenopausal at both visits (RR: 2.26; 95% CI: 1.31-3.90).

**Table 4.** Transition to high-risk category by visit 1 Lp(a) and menopausal transition status. Values expressed as percent of individuals whose Lp(a) increased to ≥125 nmol/L (95% CI). n represents the number of women who transitioned to ≥125 nmol/L in that subgroup.

|  | Overall | <75 nmol/L at visit 1 | 75-125 nmol/L at visit 1 |
| --- | --- | --- | --- |
|  | **% that increased to ≥125 nmol/L at visit 2** |  |  |
| **Women who underwent menopause** | 6.0<br>(3.9, 8.8)<br>(n=25) | 0.9<br>(0.2, 2.5)<br>(n=3) | 56.4<br>(41.0, 70.7)<br>(n=22) |
| **Women who remained pre-menopausal** | 4.0<br>(2.5, 6.0)<br>(n=21) | 0.2<br>(0.01, 1.3)<br>(n=1) | 29.0<br>(19.6, 40.6)<br>(n=20) |
| **Women who remained post-menopausal** | 2.6<br>(2.1, 3.1)<br>(n=93) | 0.1<br>(0.04, 0.4)<br>(n=4) | 28.0<br>(23.3, 33.2)<br>(n=89) |

Among women with Lp(a) <125 nmol/L at visit 1, the probability of having Lp(a) ≥125 nmol/L at visit 2 increased with higher visit 1 Lp(a) values (Figure 1). Across the full cohort, the prevalence of elevated Lp(a) increased for all groups between visits, but the largest absolute increase occurred among women who transitioned through menopause, from 6.8% at visit 1 to 11.8% at visit 2 (Table 5). Findings were similar when using 105 nmol/L as the risk-enhancing threshold (Supplemental Table).

**Figure 1.**
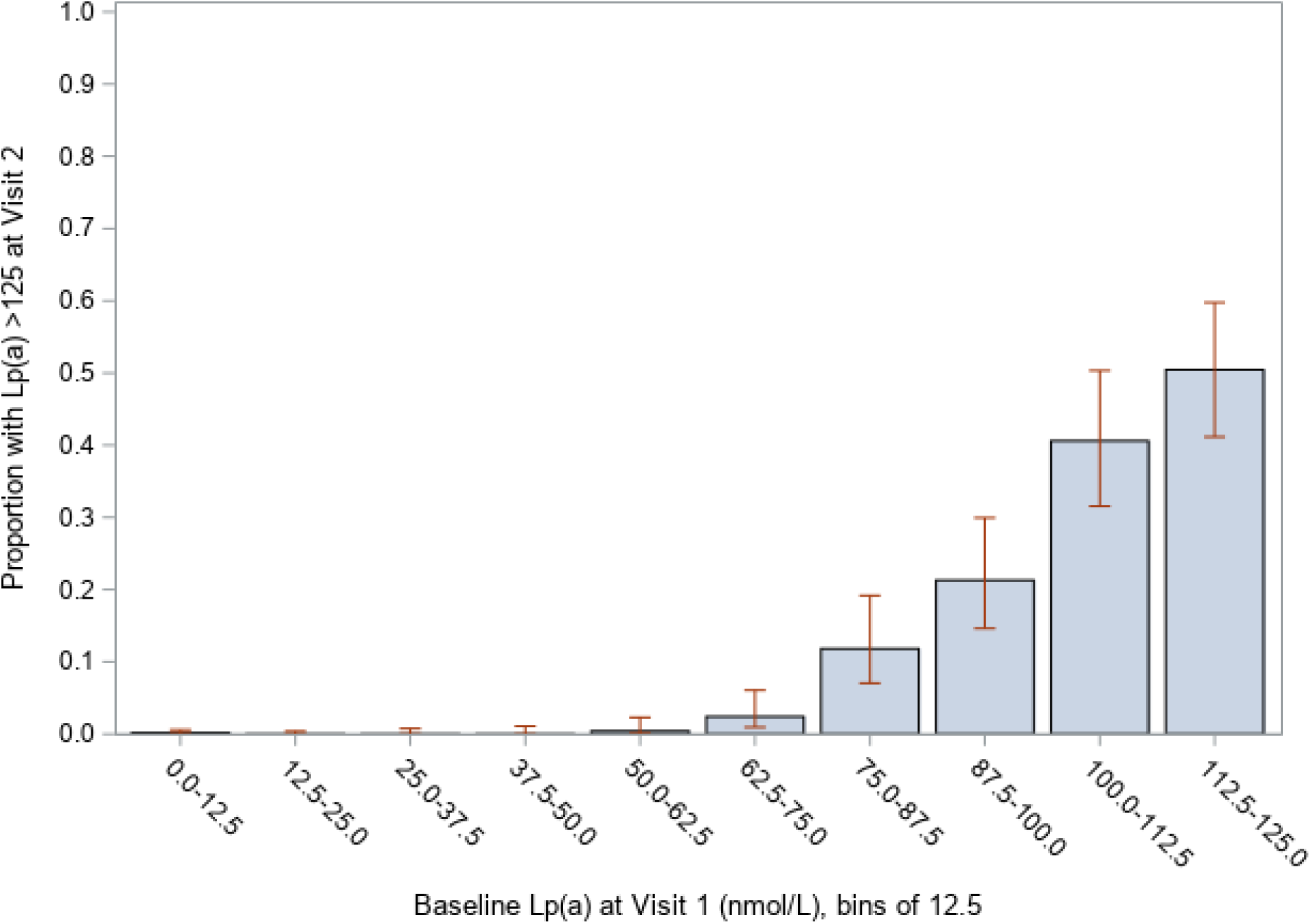
**Proportion of women who exceeded Lp(a) 125 nmol/L at visit 2 by baseline Lp(a)**

**Table 5.** Prevalence of elevated Lp(a) (. ≥**125 nmol/L) at visit 1 and visit 2 by menopausal transition status.**

|  | **% with elevated**<br>**Lp(a) at visit 1** | **% with elevated**<br>**Lp(a) at visit 2** |
| --- | --- | --- |
| **Overall** | 9.4<br>(n=428) | 10.7<br>(n=487) |
| **Women who underwent**<br>**menopause** | 6.8<br>(n=28) | 11.8<br>(n=49) |
| **Women who remained**<br>**pre-menopausal** | 6.8<br>(n=36) | 9.0<br>(n=48) |
| **Women who remained**<br>**post-menopausal** | 10.1<br>(n=364) | 10.8<br>(n=390) |

## Discussion

In this large longitudinal cohort, the menopausal transition was associated with greater increases in Lp(a) than those observed in women who remained pre- or postmenopausal, particularly among women with intermediate baseline levels. Although absolute changes were modest overall, among women with intermediate baseline levels, women who transitioned through menopause experienced median increases approximately fourfold higher than those in the comparison groups. These larger absolute increases translated into a substantially higher likelihood of crossing established ASCVD risk-enhancing thresholds, even after adjustment for age. Findings suggest that the menopausal transition may represent a clinically important window for repeat Lp(a) testing, especially in women with intermediate levels of Lp(a) prior to menopause.

These findings collectively challenge the widespread clinical perception that Lp(a) is relatively stable across the lifetime because it is genetically determined. They also are consistent with an analysis of temporal stability of Lp(a) in the UK Biobank^24^ as well as a recent analysis in the Atherosclerosis Risk in Communities (ARIC)^25^ study which found that individuals with intermediate levels of Lp(a) may benefit from repeat assessments due to fluctuations. While Lp(a) concentration is strongly heritable, accumulating evidence suggests that this assumption does not generalize to all populations or life-course stages. Menopause represents an example in which biologically meaningful changes in Lp(a) may occur, similar to observations in individuals with chronic kidney disease, liver disease, or systemic inflammatory states.^26^ Extrapolating evidence of Lp(a) stability from younger, healthier populations to menopausal women may therefore obscure clinically relevant change and delay appropriate screening or intervention in groups for whom Lp(a) is not static.

The clinical implications are notable. Approximately 6% of women who underwent menopause between study visits developed elevated postmenopausal Lp(a) levels that would not have been detected by a single premenopausal measurement. Among women with intermediate visit 1 Lp(a), more than half later exceeded 125 nmol/L and would have been missed by a one-time premenopausal test. These results provide empirical support for emerging recommendations advocating repeat Lp(a) assessment after menopause, particularly for women with intermediate levels who may be most likely to cross risk-enhancing thresholds. Notably, the observed patterns were consistent when applying a lower risk-enhancing threshold of 105 nmol/L, a cut point increasingly supported by recent European and international consensus statements. This suggests that menopause-related reclassification remains clinically relevant even as definitions of elevated Lp(a) evolve, reinforcing the importance of reassessment during this period. These findings do, however, underscore uncertainty regarding which aspects of Lp(a) exposure drive ASCVD risk in women. Whether risk reflects long-term premenopausal exposure, a new postmenopausal steady state, or the dynamic increase accompanying menopause remains unclear. Assumptions of lifelong Lp(a) stability may impede efforts to disentangle these distinct risk pathways.

Our findings are consistent with prior work from the Copenhagen General Population Study, which reported higher mean Lp(a) levels after menopause, although their study did not draw direct comparisons to women who remained pre- and post-menopausal.^6^ The study compared the life course trajectory of Lp(a) in women to that of men, reporting an additional increase around age 50. Further, the Study of Women’s Health Across the Nation evaluated changes in Lp(a) relevant to the final menstrual period and found a linear pattern consistent with age-related increases; however, differences in study design, follow-up intervals, and operationalization of the menopausal transition complicate direct comparison.^17^ The present study adds novel evidence to the literature and strengthens inference regarding the effect of menopause on Lp(a) levels by quantifying the magnitude of menopause-associated change, including absolute differences in Lp(a) and the age-adjusted risk of exceeding established ASCVD risk-enhancing thresholds.

Additional strengths of the present study include its longitudinal design and large sample size. Further, the use of an isoform insensitive immunoturbidimetry method to measure Lp(a), which is less affected by heterogeneity in apolipoprotein(a) isoforms, provides more accurate and consistent results.^27^

A limitation of the present study is that menopausal status was self-reported. Although variables capturing age at menopause and time since last period were available, these were conditional on self-reported status and did not permit precise classification beyond 12 months since last menses. Misclassification of menopausal status is therefore possible. Complete data for onset of kidney and liver disease were not available, preventing adjustment for these conditions which may also influence Lp(a).^26^ Additionally, all participants were of European descent. Future work in this area should include individuals known to have higher Lp(a) levels, including women of African and South Asian descent.^28, 29^ These women are therefore more likely to have elevated and intermediate levels of Lp(a), and the rate of women in these groups crossing the risk-enhancing threshold is likely to be higher. An additional limitation is that the women who returned for a second assessment were chosen via a convenience sample of individuals who lived nearby to the UK Biobank coordinating center.

In conclusion, Lp(a) increases at a higher rate during the menopausal transition, particularly among women with intermediate premenopausal levels. These findings highlight menopause as a clinically important period during which assumptions of Lp(a) stability may not hold and support repeat Lp(a) measurement after menopause to improve cardiovascular risk stratification in women.

## Data Availability

All data used in this study are available from the UK Biobank (https://www.ukbiobank.ac.uk) to qualified researchers for health-related research in the public interest. Data were accessed under an approved UK Biobank application and according to data use agreements in place between the UK Biobank and the University of North Carolina at Chapel Hill. Requests to access these data can be made directly through the UK Biobank.

## Acknowledgments & Disclosures

Funding

This study was funded by the National Heart, Lung and Blood Institute of the National Institutes of Health (T32HL129982).

## STROBE Statement—checklist of items that should be included in reports of observational studies

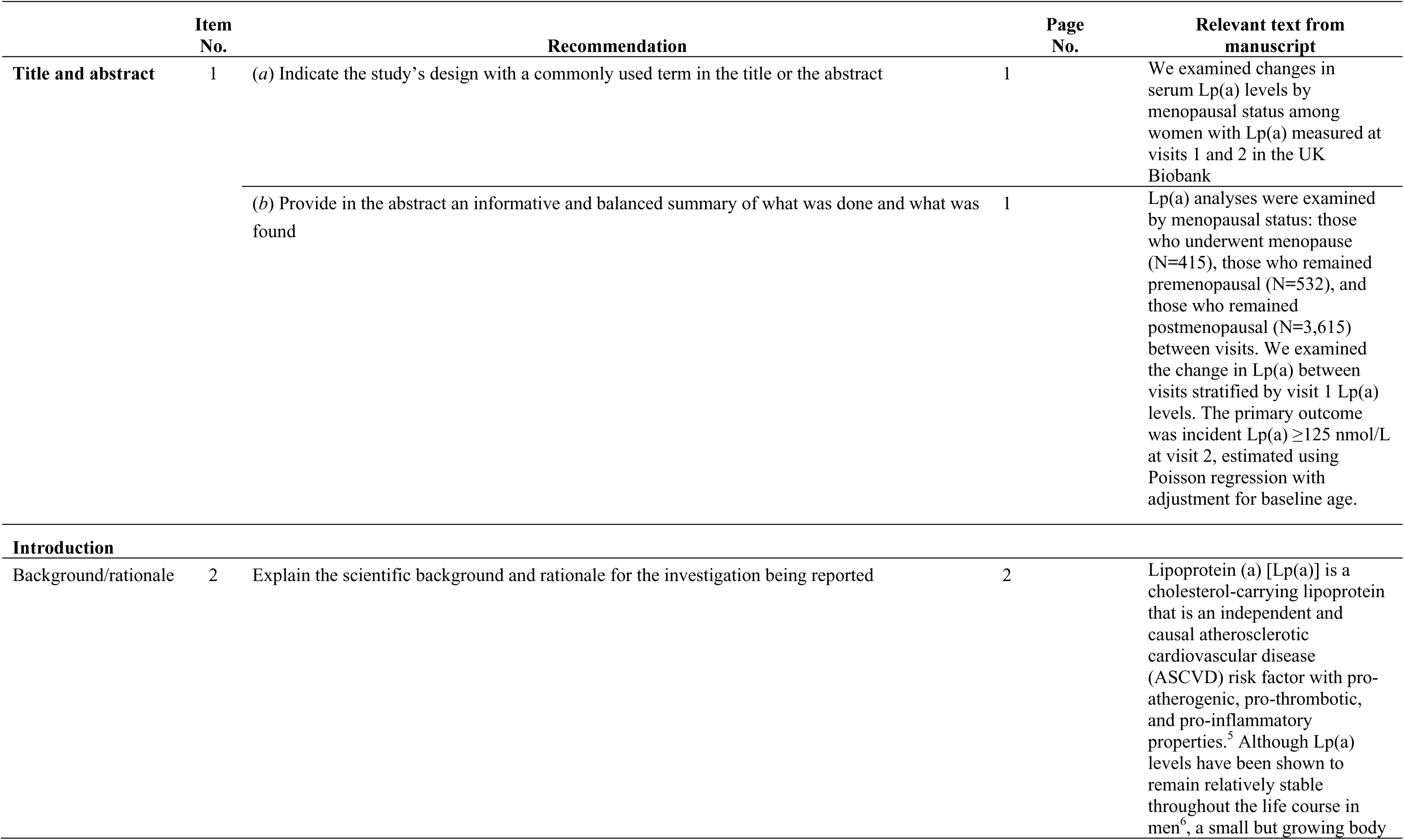
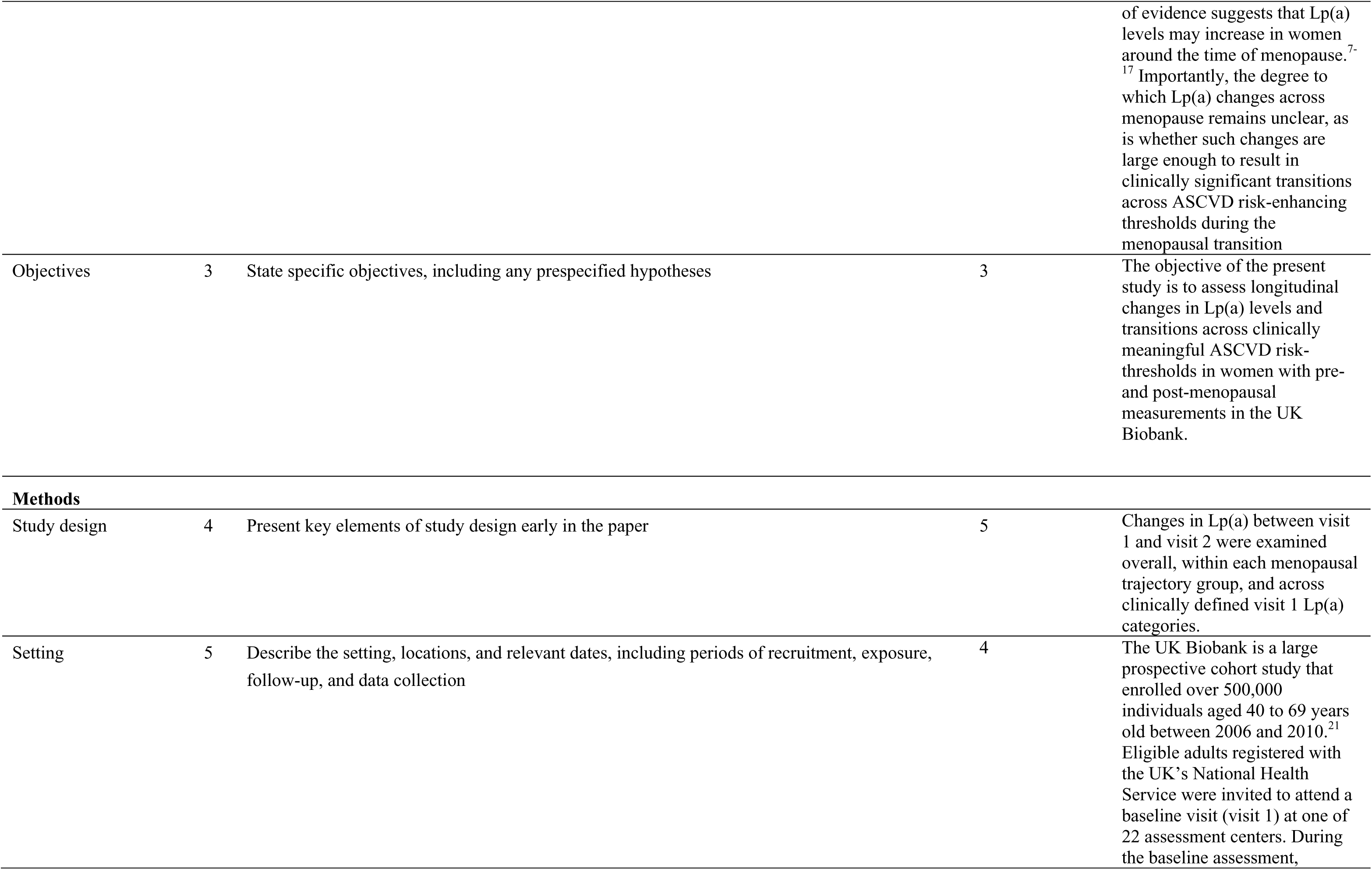
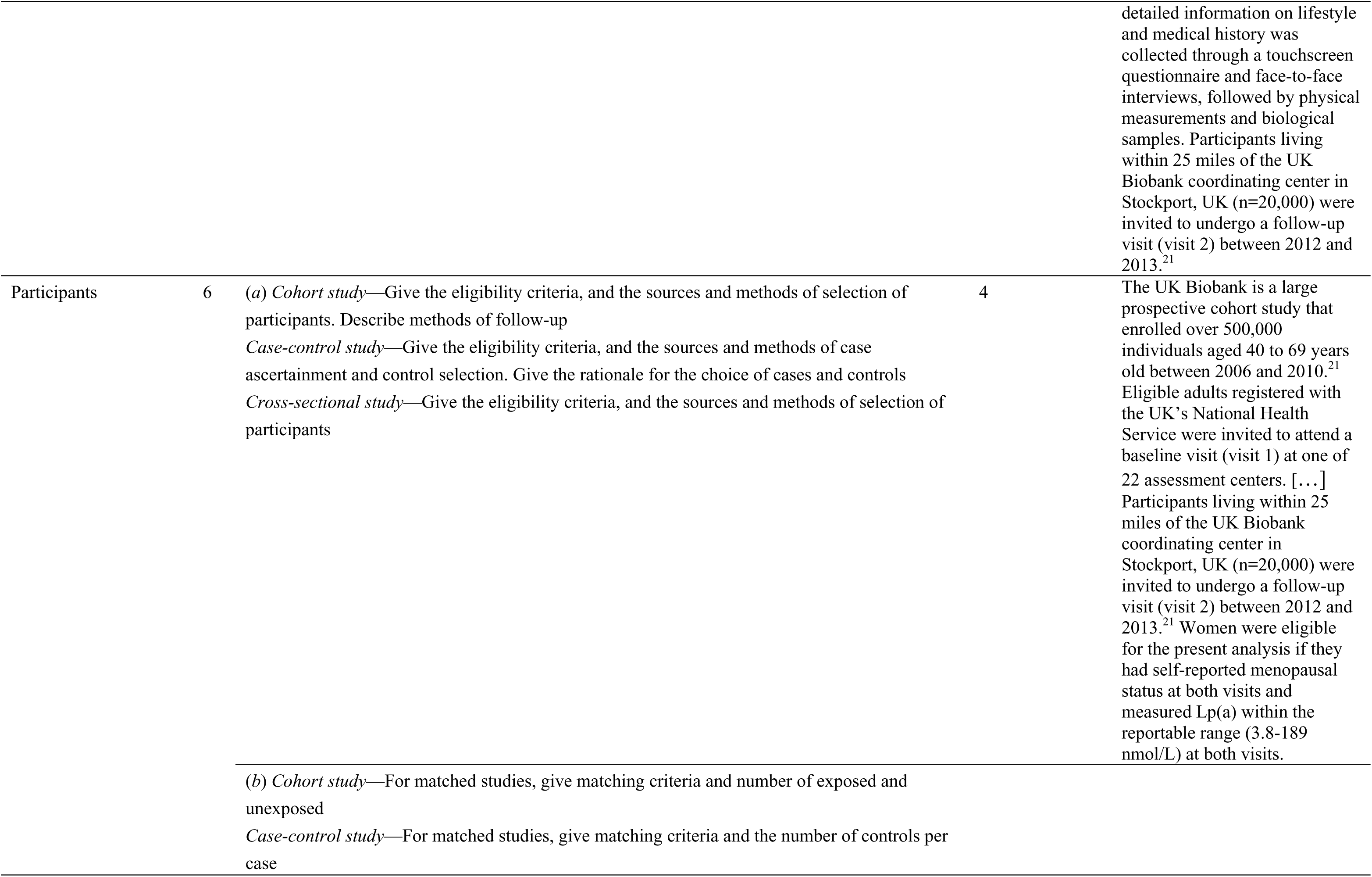
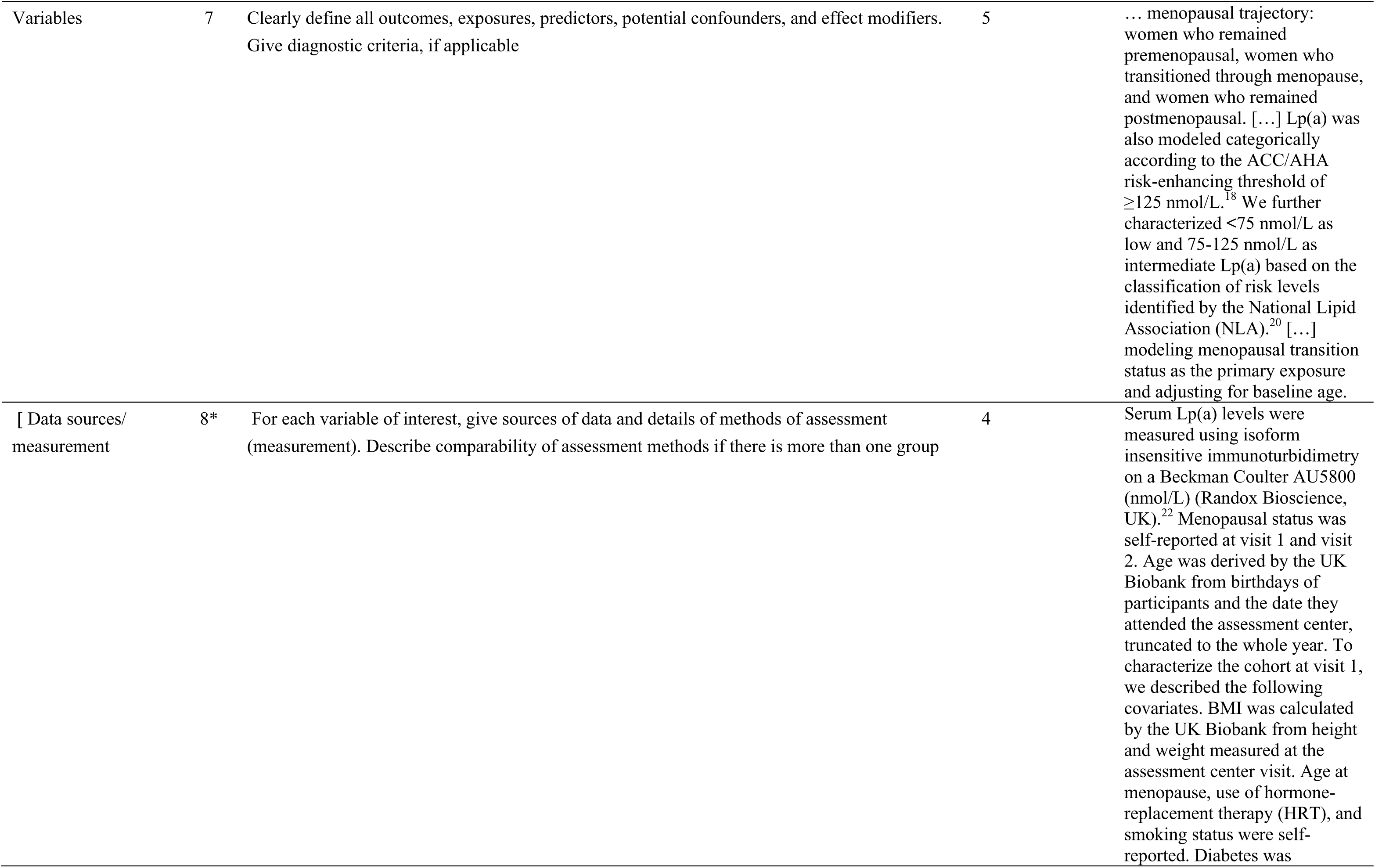
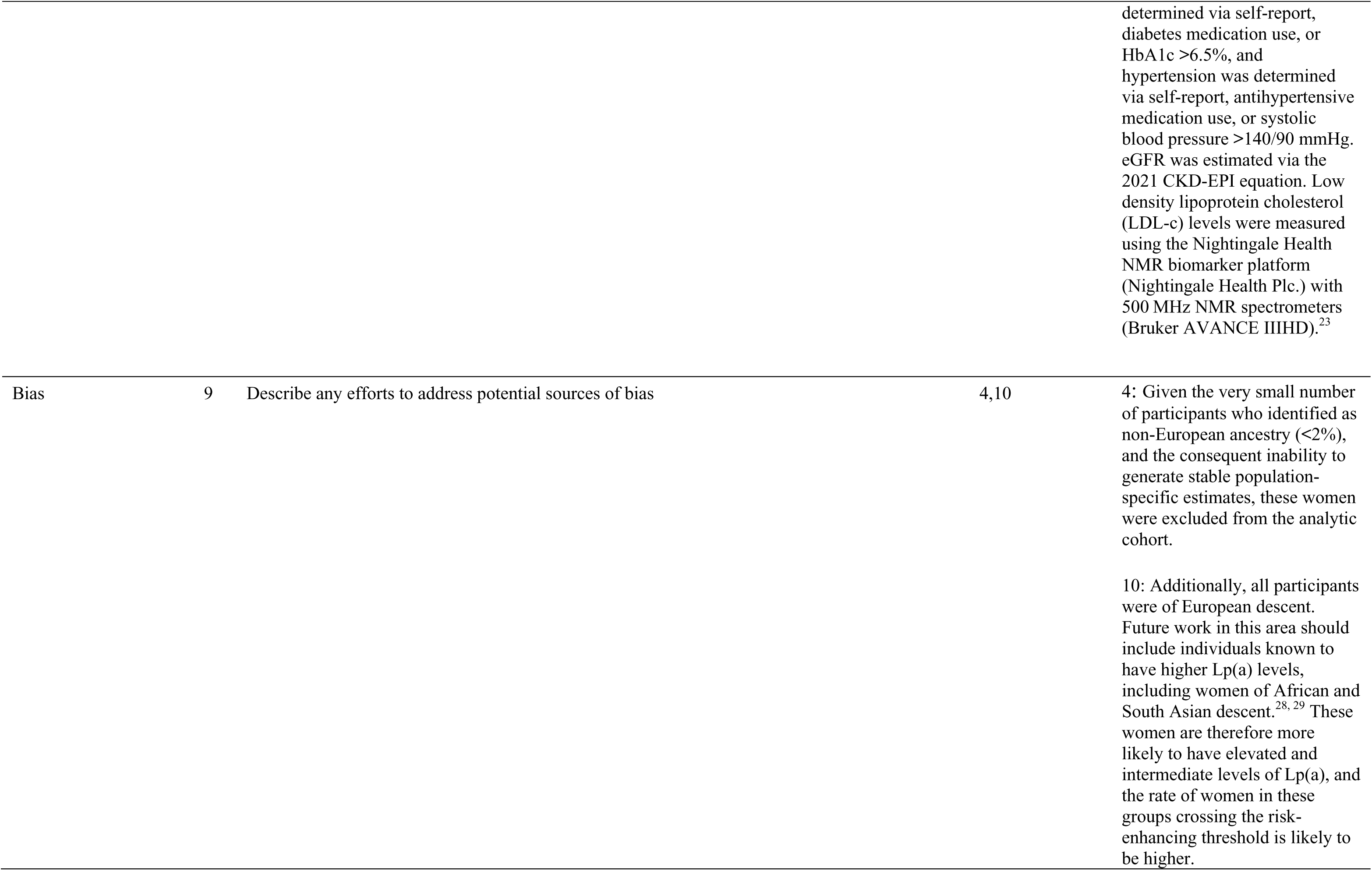
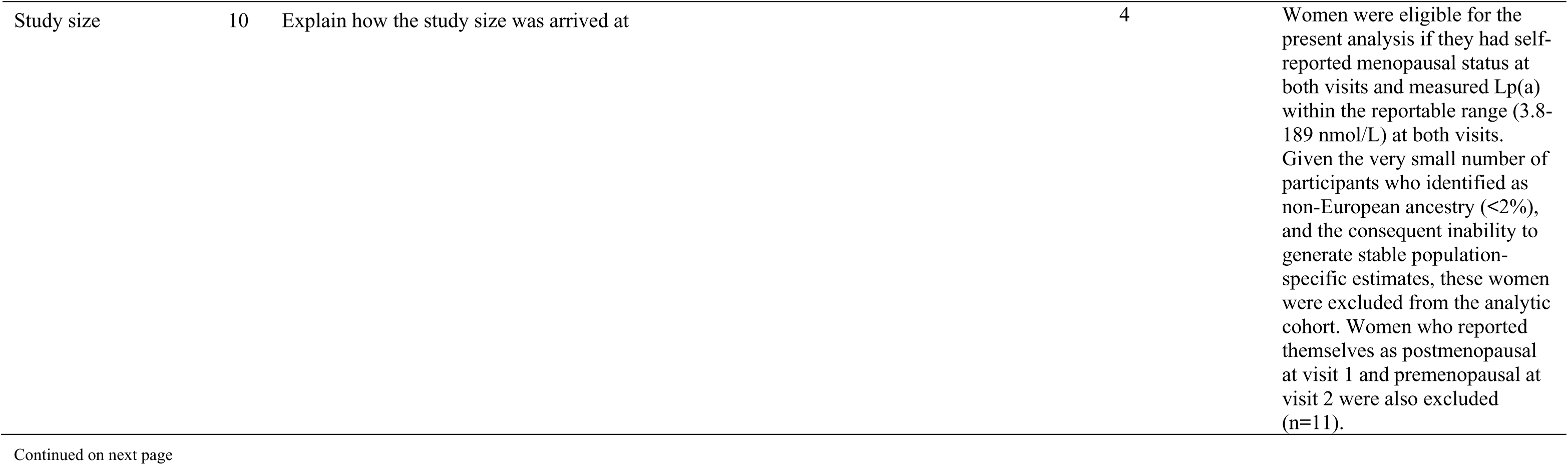
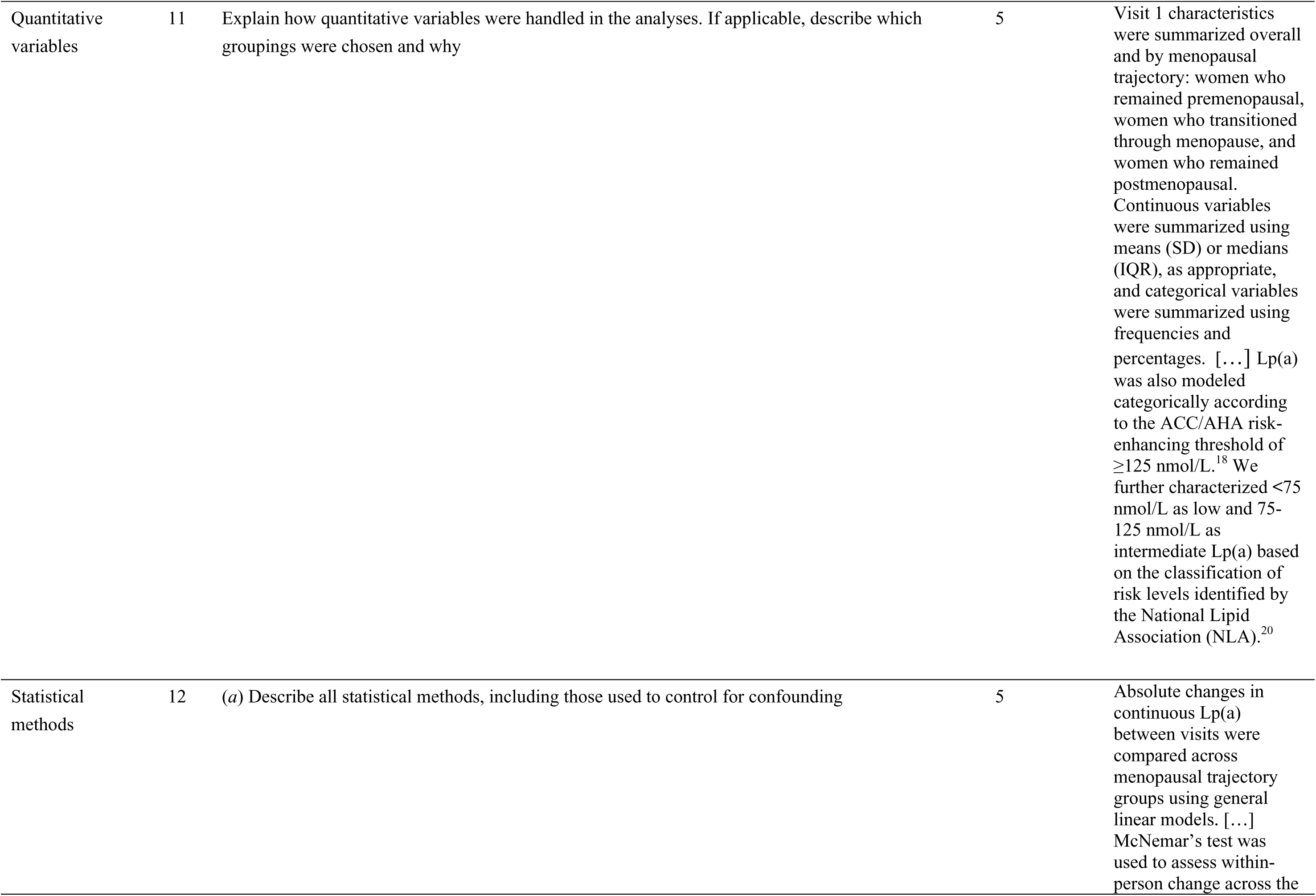
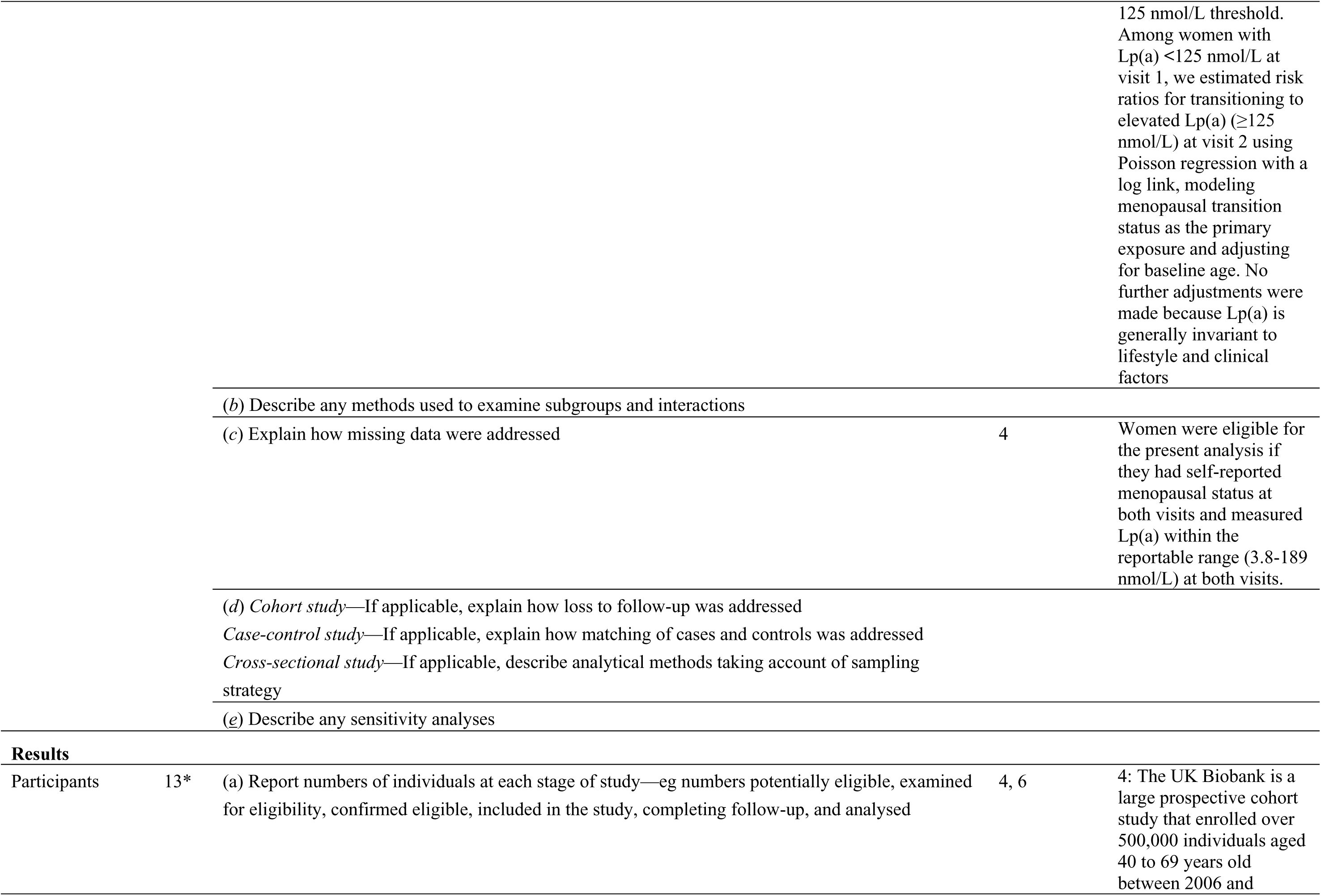
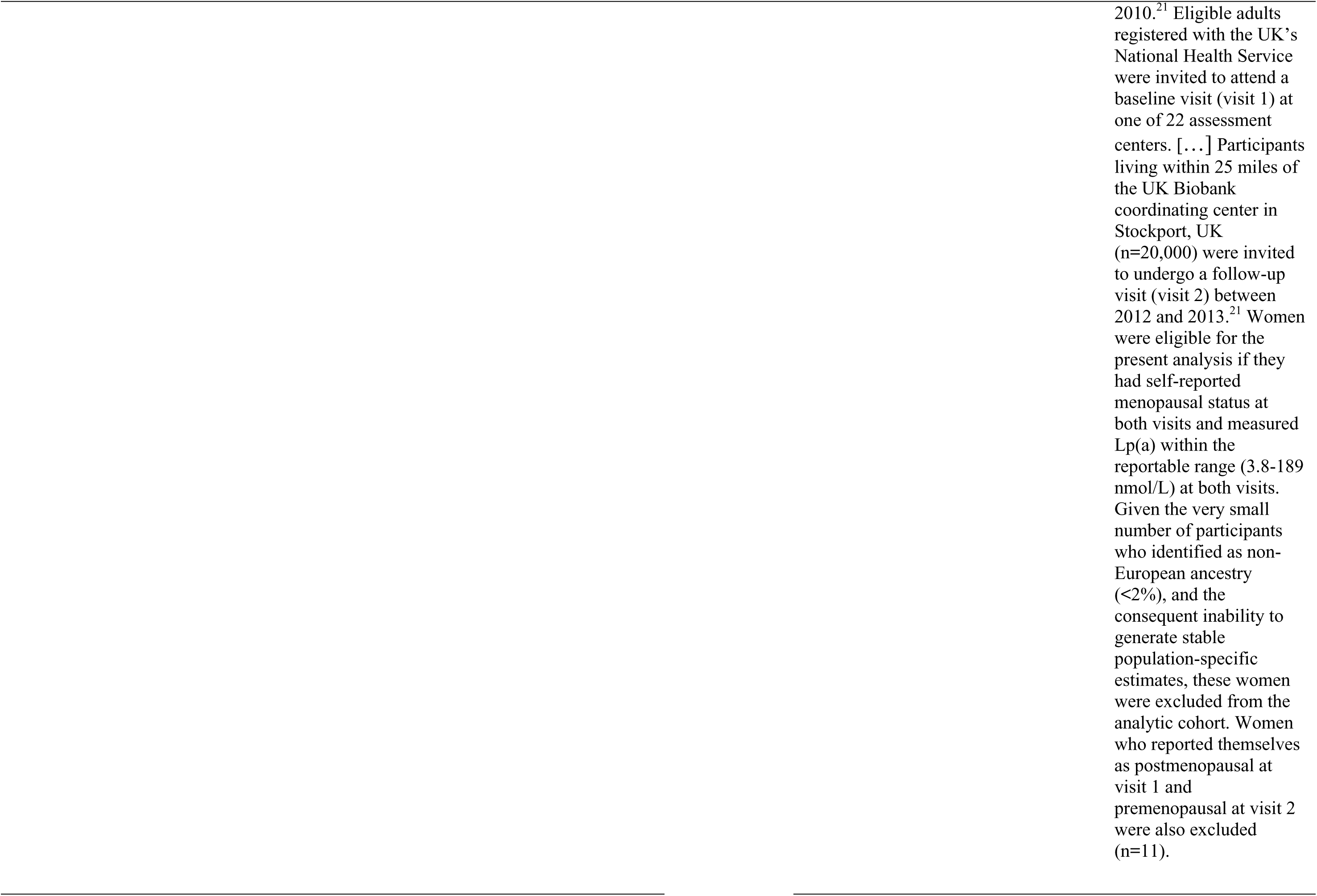
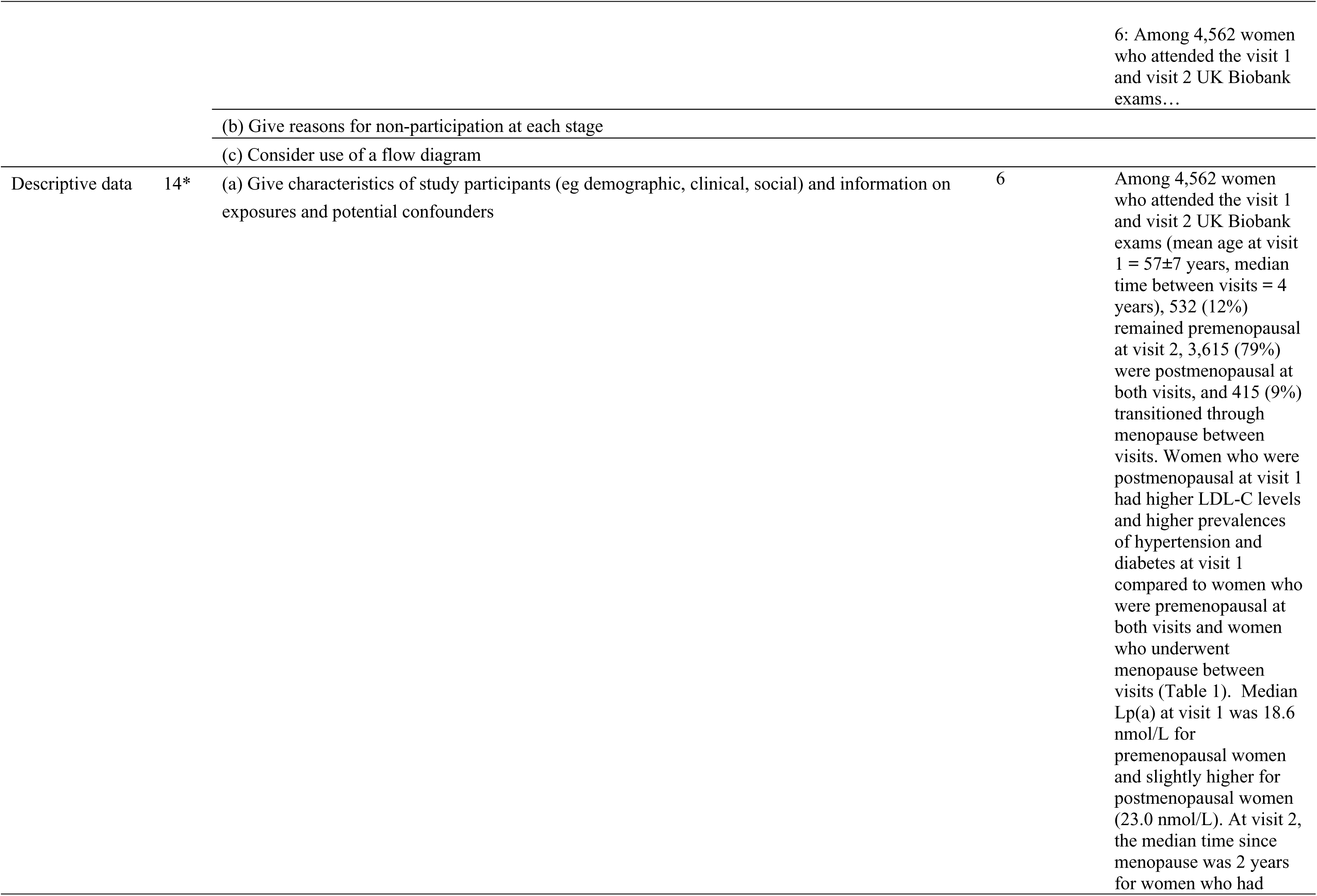
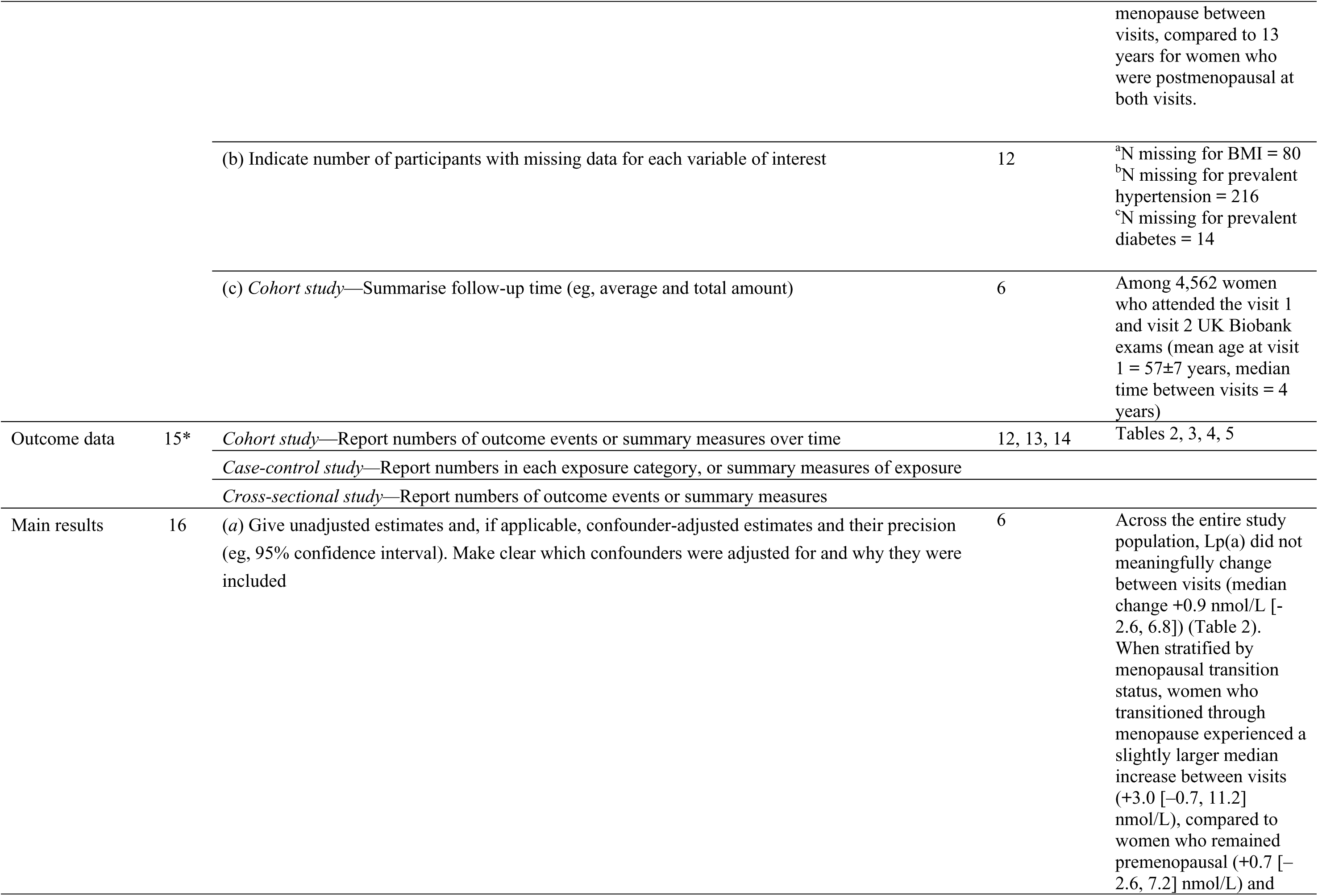
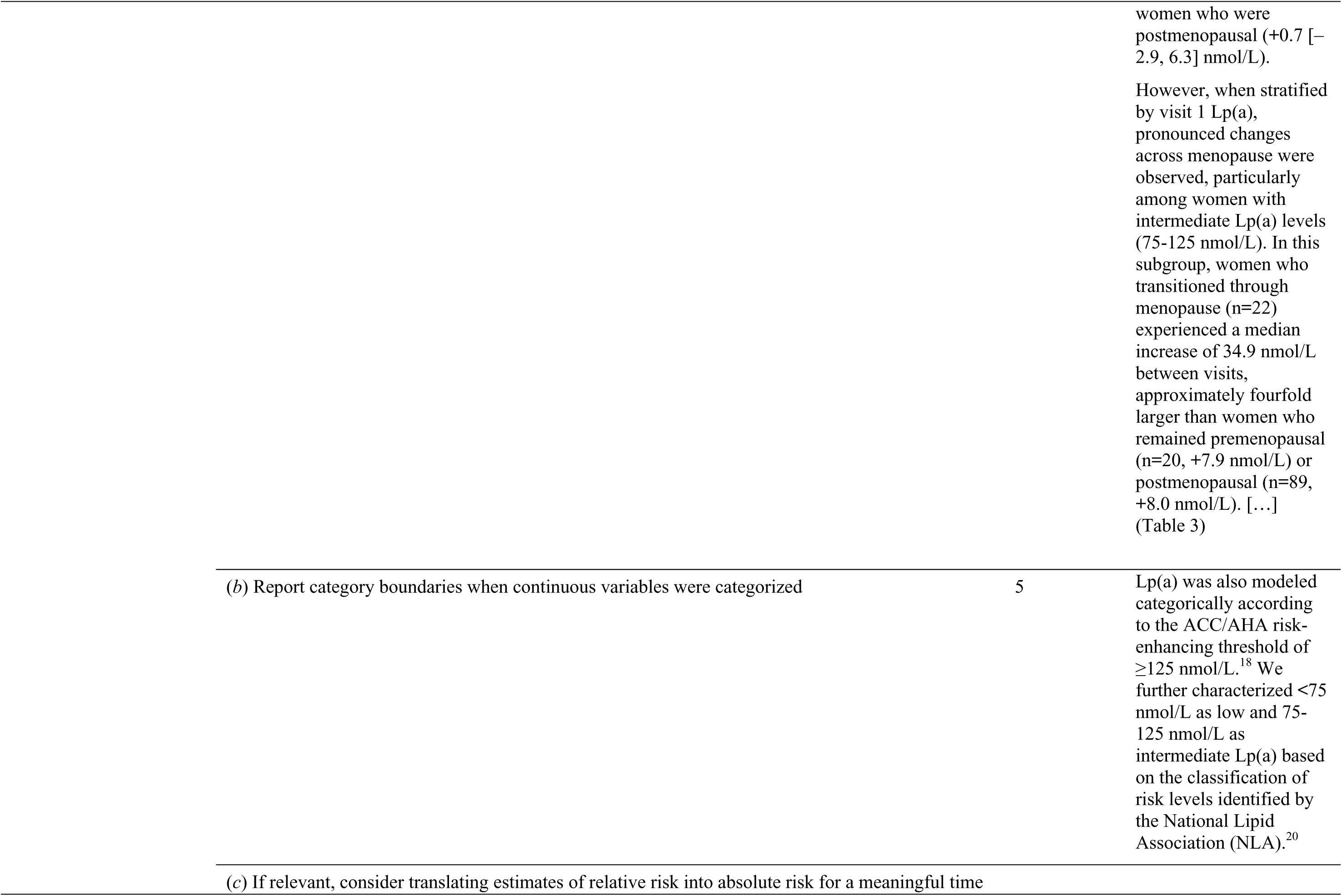
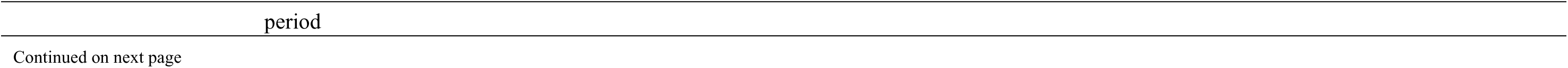
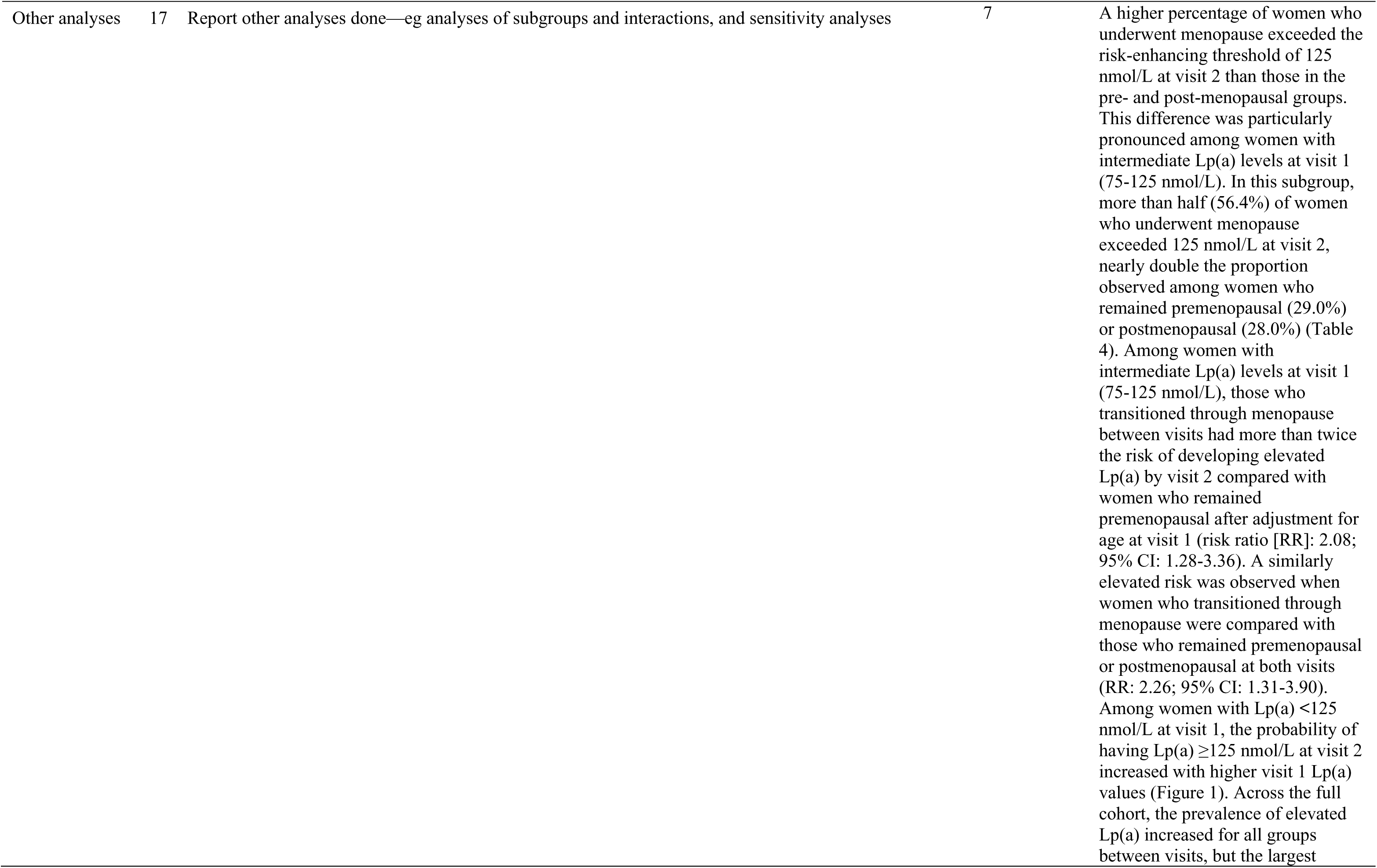
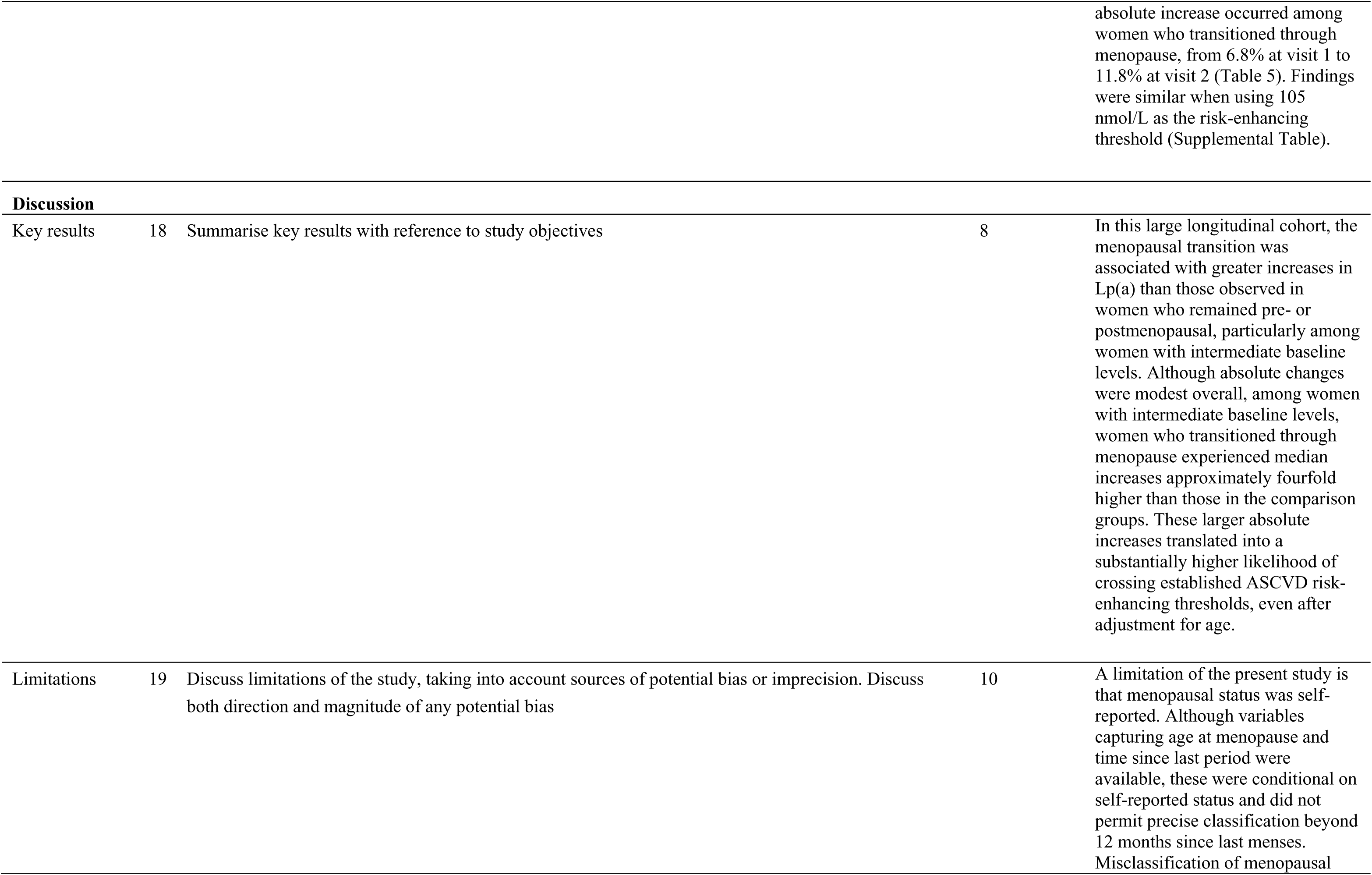
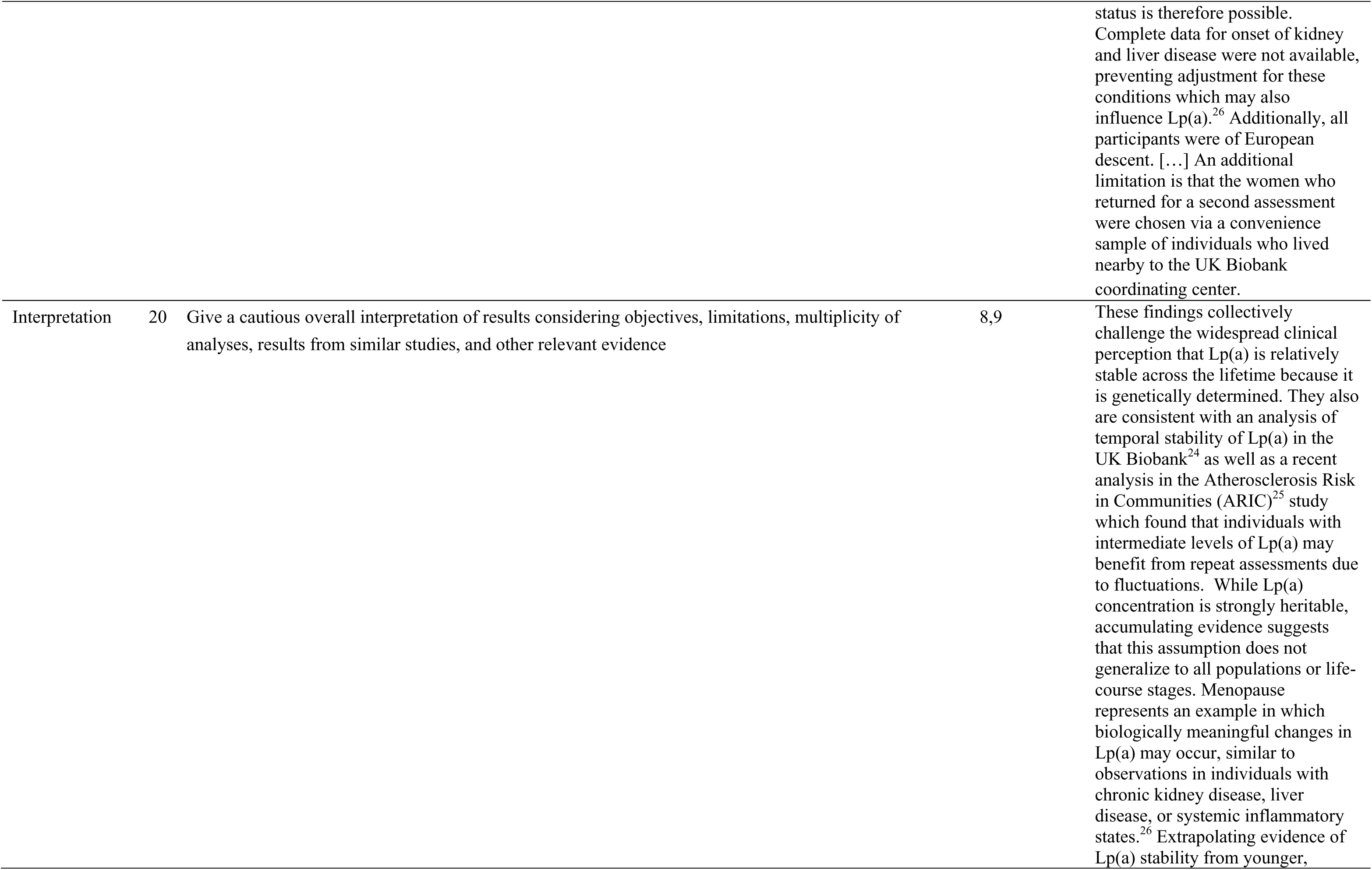
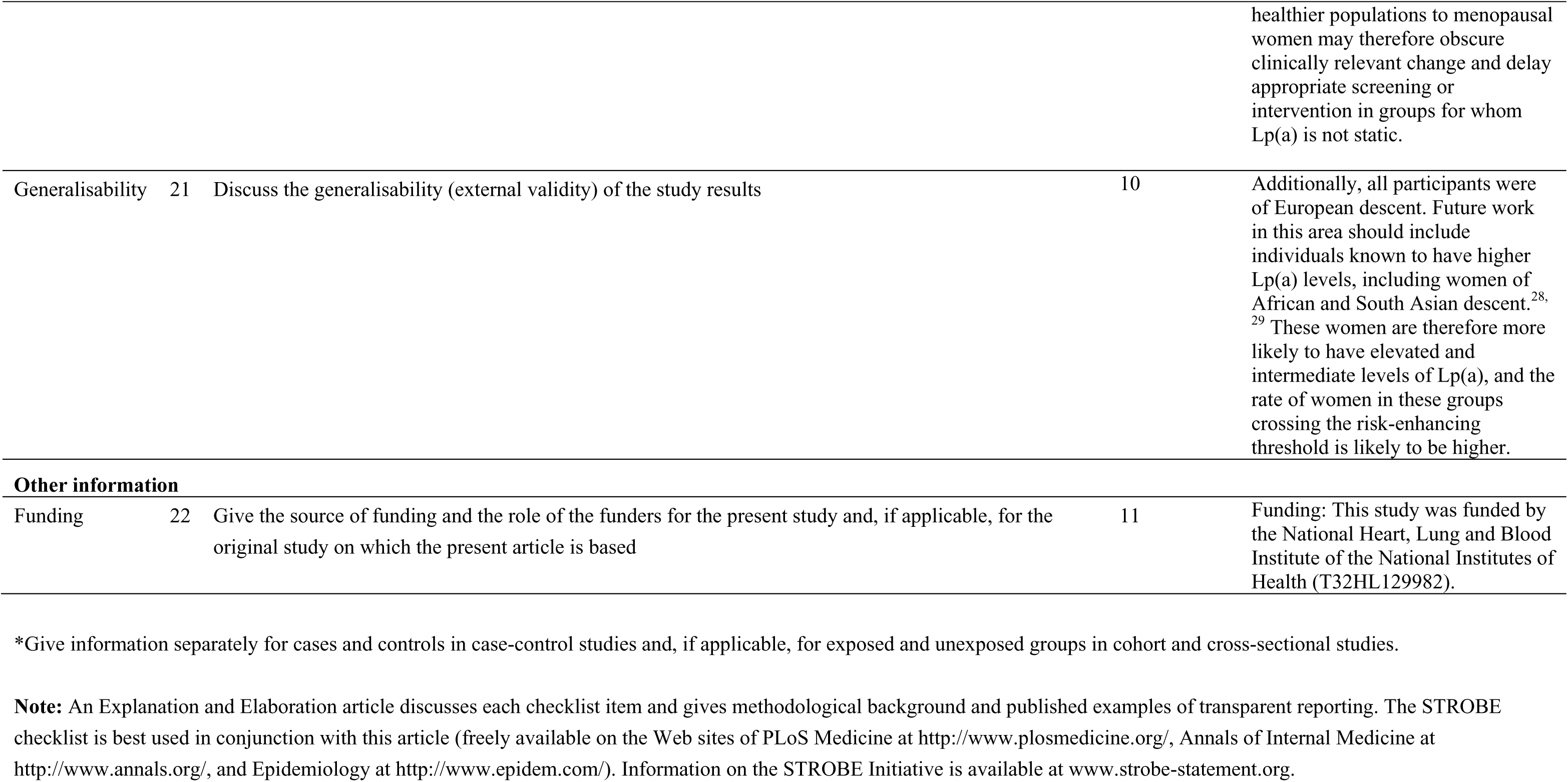

